# Light weight deep learning-based auto-quantification system for bright-filed HER2 dual in situ hybridization image analysis

**DOI:** 10.1101/2025.07.27.25331550

**Authors:** Chao-Ying Huang, Jia-Rong Lin, Po-Chi Huang, Chen-Hsuan Liao, Chu-Cheng Yen, Li-An Chu

## Abstract

The evaluation of erb-b2 receptor tyrosine kinase 2 (ERBB2 or HER2) gene amplification status through Dual in Situ Hybridization (DISH) currently relies on manual assessment by pathologists. There are several deep learning-based algorithms for H&E or ISH analysis. However, DISH analysis tools are still lacking.

We developed a fully automated deep learning-based quantification system to assist pathologists in identifying the most relevant cells throughout the entire DISH image. In the comparison between pathologists and the auto-quantification system, the overall percentage agreement (OPA) by case was 88. 9% (80/90).

These results demonstrate that each image, with a processing time of approximately 1 minute, achieves similar results compared to pathologists’ assessments, while the manual procedure will take 10-20 times longer to examine the same specimen.

This approach offers a versatile system for bright-field HER2 DISH image analysis. The system provides faster, cheaper, standardized, and versatile diagnostic tools to aid pathologists in the HER2 DISH diagnostic process.

## Introduction

The increasing incidence of breast cancer, particularly among younger individuals, has made it the most diagnosed malignancy in women globally (^1^). Overexpression and amplification of human erb-b2 receptor tyrosine kinase 2 (ERBB2 or HER2), a member of the epidermal growth factor receptor family, are observed in 15–20% of breast cancer cases (^2^;^3^) and are frequently associated with a higher risk of recurrence and mortality (^4^;^5^). In recent years, first-line treatment options have predominantly included chemotherapy in combination with HER2-targeting agents such as trastuzumab (Herceptin) and pertuzumab (Perjeta), alongside the mitotic inhibitor taxane (THP) (^6^;^7^). Research has shown that early identification and intervention before the spread of malignant can significantly reduce mortality (^8^). This underscores the critical importance of accurately determining HER2 positivity, either through gene amplification or protein overexpression, to classify the breast cancer sub-type and ensure that patients receive the most appropriate HER2-targeted therapy, thereby optimizing their chances of survival.

Current practice for determining HER2 status involves immunohistochemistry or in situ hybridization techniques, including but not limited to fluorescence in situ hybridization (FISH), chromogenic in situ hybridization (CISH), and dual in situ hybridization (DISH). These procedures require pathologists to manually select 20 cells, record signals, and calculate ratios from the in-situ hybridization datasets—a process that is time-consuming, labor-intensive, and inconsistent due to the inability to inspect every cell within the entire image and different standards of individual pathologist. Moreover, with the increasing demand for new tests in various fields, the growing responsibilities of pathologists, and the shortage of new pathologists entering the profession (^9^), there is an urgent need for a system that can expedite the quantification process.

The advantages of using DISH over FISH or CISH include improved visual clarity, easier interpretation, and particular effectiveness for cases with borderline HER2 expression. Both DISH and CISH samples can be stored for longer periods, whereas FISH samples are susceptible to severe photobleaching. Additionally, brightfield microscopy, used for DISH and CISH, is more standardized and widely available compared to the specialized fluorescent microscopy required for FISH imaging. When directly comparing DISH to CISH, DISH often provides better image contrast, which is also crucial for accurate diagnosis.

Significant progress has been made in the automated analysis of FISH and CISH images, with previous studies reporting a high correlation of 0.98 with manual HER2/Chromosome 17 (Chr17 or CEP17) ratios (^10^). While mixed manual and algorithmic approaches for DISH images have achieved impressive Overall Percentage Agreement (OPA) rates as high as 98.8% (^11^), these methods still require extensive manual intervention and have yet to demonstrate consistent performance across varying specimen conditions. Our raw image dataset exhibits considerable variation due to differences in each step when staining and digitalizing the specimens (1). While these variations don’t seem to affect pathologists to perform the diagnosis, previous research on DISH image analysis relies on manual annotation on individual cell body, which required additional redundant work that are not practical in clinical practice (^12^).

We propose a HER2DISH image auto-quantification system designed to assist pathologists in assessing breast cancer cells stained with methods such as, but not limited to, the VENTANA HER2 Dual ISH DNA Probe Cocktail. Our system leverages deep learning methods for Her2/Chr17 and cell border segmentation, having higher resilience towards image variation and resulting in increased accuracy. Additionally, the system demonstrates high accuracy and flexibility, allowing for manual correction of results. Ultimately, this auto-quantification system is sufficiently robust to support pathologists in DISH image analysis.

## Results

Currently, the manual process requires an experienced pathologist to first assess the presence of any clusters exhibiting HER2 amplification. If such clusters are identified, cells for quantification are selected from within these clusters; otherwise, cells are chosen from areas free of noise and artifacts. An initial set of 20 cells is then selected from the designated area, ensuring that the cells are of average size. Preference is given to cells exhibiting signal levels close to the highest average observed. Following the selection of these 20 cells, and potentially an additional 20 if needed, the final determining metrics for HER2 status are computed: the HER2/Chr17 ratio (i.e., the ratio of HER2 signals to Chr17 signals across all cells) and the average HER2 signal per cell. Metrics are then evaluated against standards aligned with the 2023 HER2 Breast Cancer Testing Guidelines established by the American Society of Clinical Oncology and the College of American Pathologists (ASCO/CAP) (^13^).

Our auto-quantification system mirrors the manual workflow (2), starting with the deep learning based automated segmentation of HER2, Chr17, and cell borders for all cells in the specimen. Utilizing the HER2 signal mask, the system identifies heat zones indicative of HER2 gene expression, enabling the selection of cells from amplification/overexpression clusters. The system then automated calculated and assigns a ranking number to each segmented and quantified cell using a proposed algorithm that evaluates individual cells based on various parameters, including but not limited to sphericity, area, and HER2/Chr17 ratios. Finally, the cells are ranked by their rankings in descending order and presented to the pathologist for final adjustments. Once an adequate number of cells have been selected, the system provides results in accordance with the algorithm specified by ASCO/CAP.

### Her2 and Chr17 Signal Segmentation

The semi-supervised pixel-wise random forest classifier was selected for the segmentation process because HER2 and Chr17 exhibit distinct shapes and RGB values compared to other features in DISH images. Additionally, to enable deployment across various clinical institutions, this lightweight approach will minimize system requirements to facilitate more cost-effective deployment. For classifier training, 12 images with distinctive characteristics were combined into a large composite image to train a versatile classifier capable of handling diverse DISH images. The results indicate that the classifier is effective in recognizing HER2 and Chr17 signals.

However, we observed that in some instances, the HER2 and Chr17 signals overlap (3), leading to reduced segmentation performance. Another phenomenon observed is the formation of clusters of HER2 signals within cells when HER2 is overexpressed. While the classifier segments the cluster as a single entity, individual HER2 signals are often scattered around the cluster. Due to the strong expression of HER2 signal in this type of specimen, this type of distribution does not affect the final HER2 status classification (3). In this study, all classifiers were trained on a system featuring an 11th Gen Intel Core i7-1165G7 @ 2.80GHz × 8, 16GB of RAM, and an NVIDIA GeForce MX350 GPU. However, the workflow is designed to function on systems without a GPU.

### Comparison of classical edge based to Deep learning method

To quantify the expression level of HER2 signal in each cell, the cell border detection and segmentation become crucial for the quantification. For cell border detection, the classical edge detection method selected for comparison involved a combination of morphology filter, adaptive threshold, distance transform, and watershed algorithms. During manual segmentation, we observed that the significant variance within our dataset necessitated different kernel radii, varying thresholds, and revealed the limitations of the watershed algorithm, particularly in its inability to accurately identify cell borders in densely populated areas.

**Figure 1.**
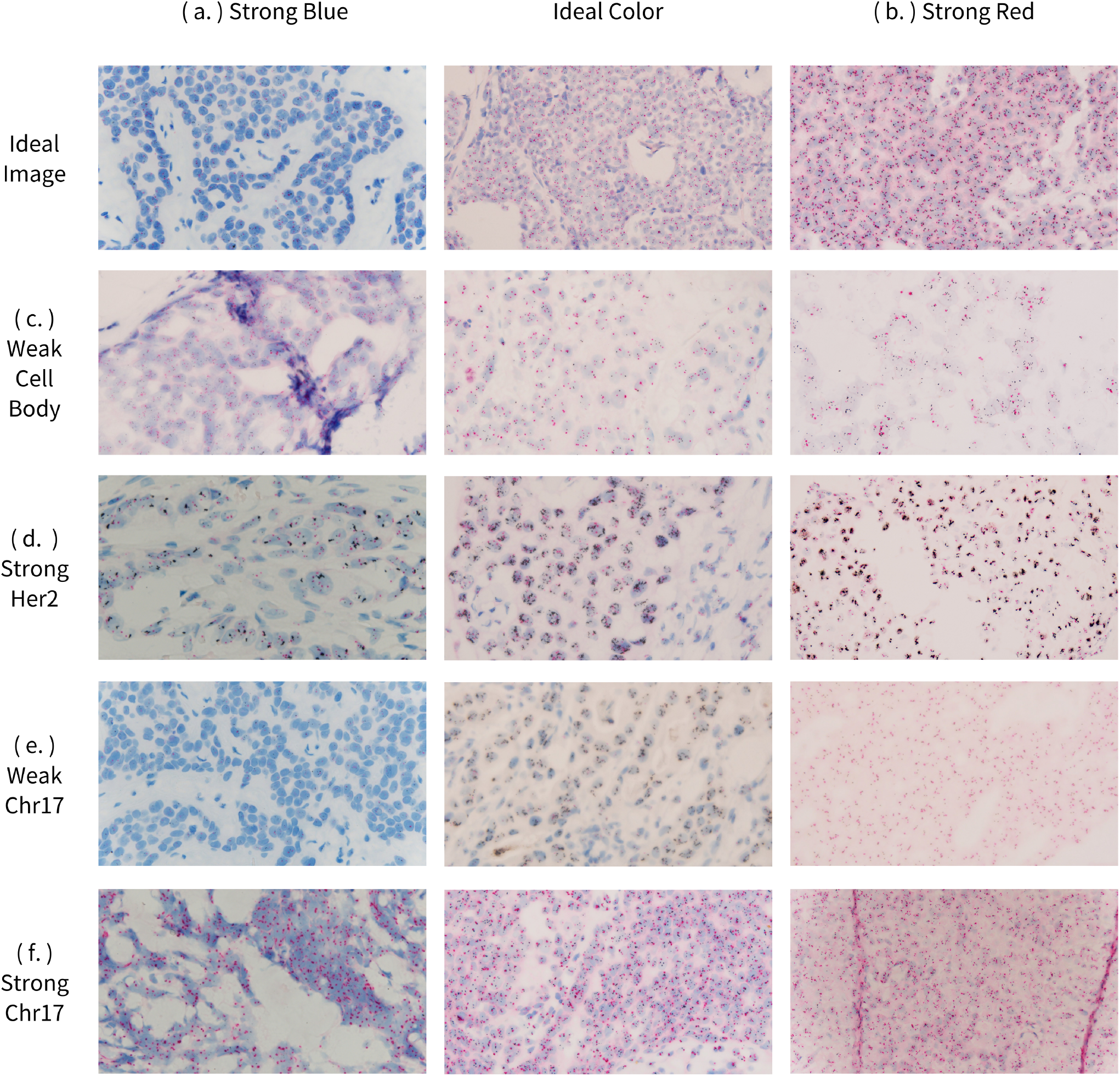
Different Staining and Imaging Results of VENTANA HER2 Dual ISH DNA Probe Cocktail Across Samples. (a.)Excessive hematoxylin staining, often due to over-staining or inadequate washing, can obscure HER2 (black) and Chr17 (red) signals. (b.)Variations in Chr17 red intensity may result from inconsistent fixation times affecting stain uptake. (c.)Under-or over-fixation can weaken cell body staining, appearing as pale signals. (d.)Amplified HER2 signals appear darker, often due to optimized fixation and tissue digestion, enhancing target visibility. (e.)Nuclear bubbling, common in thicker sections or those with excess paraffin, can displace and enlarge Chr17 signals. (f.)Prolonged or delayed fixation reduces Chr17 staining intensity, making the signal fainter and more challenging to quantify.

**Figure 2.**
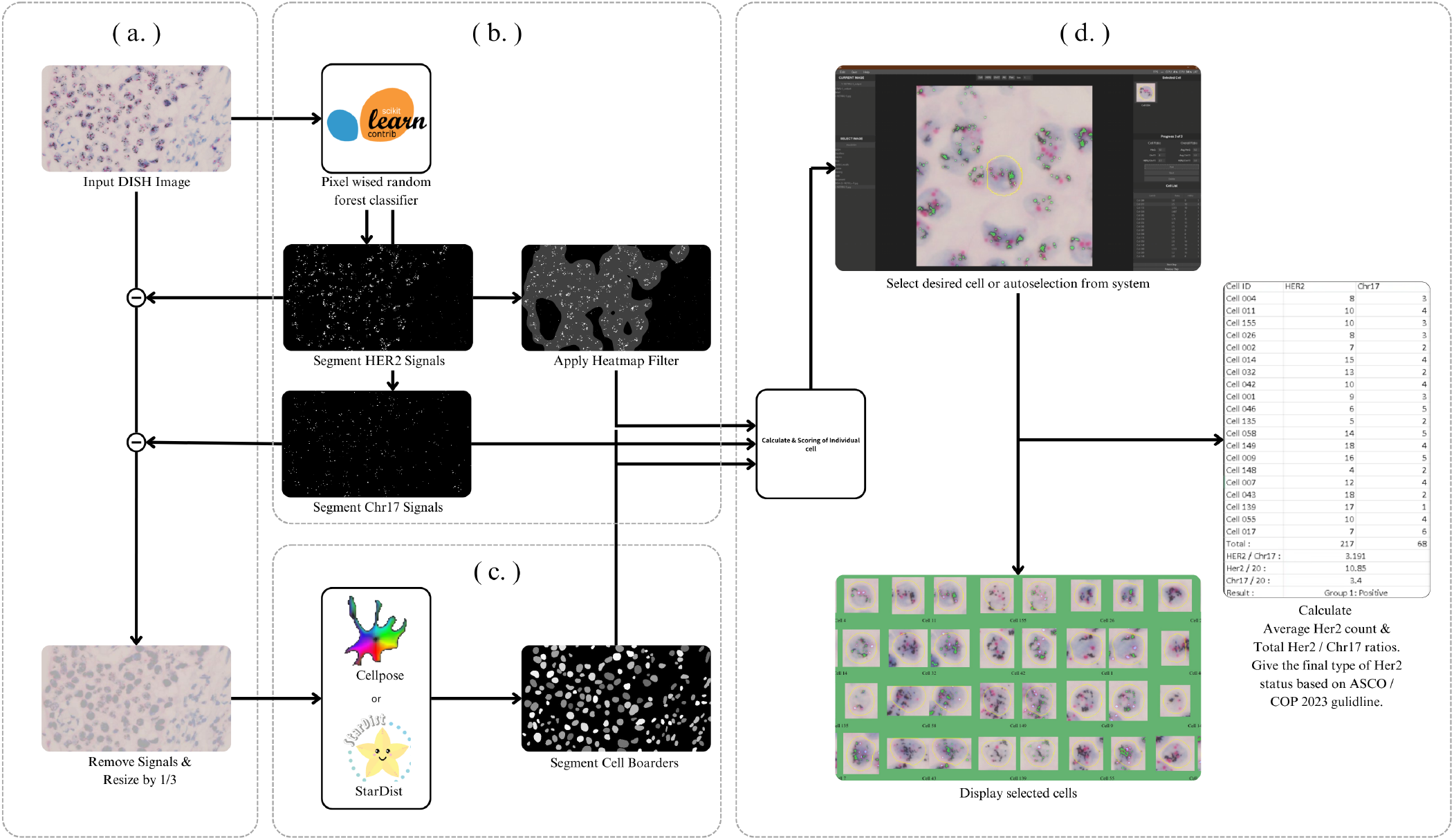
Auto-quantification system workflow. (a.) Input image into the workflow with pre-processing method of removing detected signals and pad with cell body background. (b.) Input image first get segmented by the pixel wise random forest classifier. The HER2 mask are also evaluated on the amount of the signals and apply a heat map filter to isolate areas with HER2 overexpression. (c.) Pre-processed image is then inputted into StarDist or Cellpose model to segment the cell boarders. (d.) HER2 mask, Chr17 mask, and cell boarder mask are paired up to calculate signals of individual cell and rank cells by a given score determined by different parameters. 20 cells are then selected either manually selected by pathologist or automatically by the system, results of each cells are then saved as pictures and individual stats.

**Figure 3.**
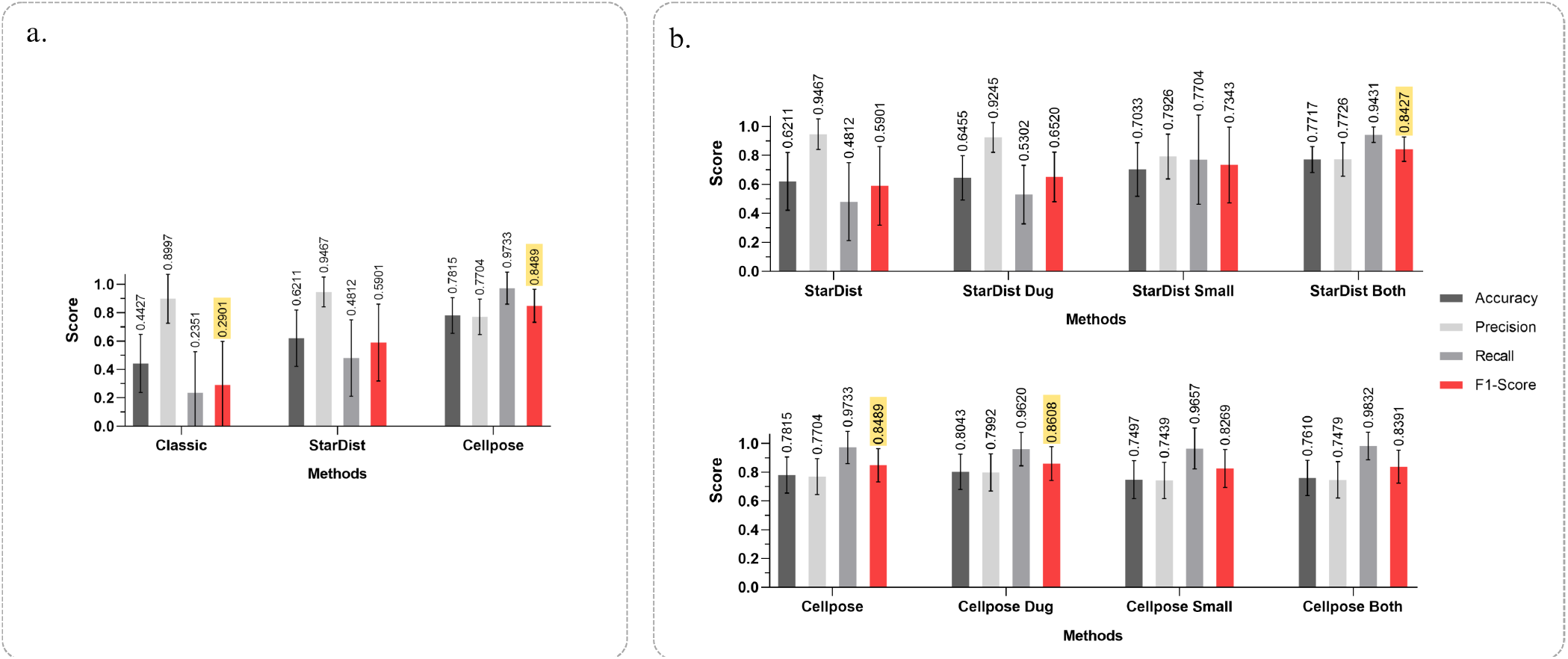
HER2 and Chr17 signal segmentation results All stats are evaluated over n = 450 cell ROI (a.) Performance comparison of classical with deep-learning methods(classical method achieve average accuracy and F1-Score of 0.4427 and 0.2901, compared to Cellpose model’s 0.7815 and 0.8489) (b.) Performance improvement of pre-processing methods with based StarDist model average accuracy and F1-Score of 0.6211 and 0.5901 compared to pre-processed StarDist’s 0.7717 and 0.8427. Comparison of StarDist and Cellpose model, StarDist with all pre-processing methods achieved average accuracy and F1-Score of 0.7717 and 0.8427, compared to Cellpose model’s without any pre-processing of 0.7815 and 0.8489, and Cellpose with signals removed of 0.8043 and 0.8608.

To overcome this problem, two deep learning approaches were applied into our test, StarDist and Cellpose models (^14^:^15^). Both algorithms were previously developed for cell detection and segmentation in microscopy images.

We tested all three approaches and found that the variability contributed to the wide range of metrics of classical edge detection approach was severe compare to the StarDist and Cellpose models (4). However, the performance of both models remains far from matching the results of manual selection. A possible reason is that these models were originally designed for cell detection in monochromatic images, where cell signals typically do not overlap with HER2 and Chr17 signals, as they do in DISH images.

### Image impanting prior to cell border detection

To mimic cell morphology in microscopy of the monochromatic image, we removed the Her2/ChR17 signals from the cells in the original DISH image (the Dug pre-processing method) and significantly improved the performance of both StarDist and Cellpose models. The most notable performance enhancement was observed in the StarDist model, where the mean F1 score increased from 0.5901 to 0.6520, and the standard deviation of the F1 score improved from 0.356 to 0.241. We attribute this performance boost to the successful removal of strong signals that were previously misidentified as cell borders (4). (you need a new figure indicating how signal removal been done)

With the Dug pre-processing method, the Cellpose model’s performance on cell segmentation also increased slightly in average F1-Score, from 0.8489 to 0. 8608.We speculate that the reason why we find limited increase in performance on the Cellpose model may be due to the Cellpose model’s pre-processing methods and training dataset being specifically designed to handle situations where signals interfere with cell borders. Consequently, we believe that removing signals at the cell borders may ever so slightly improve the overall shape of the cell mask.

To further enhance the performance of both models, we resize the image by a factor of 0.33 on both axis prior to the segmentation process. This adjustment led to significant increases in accuracy and F1 score for the StarDist model, rising from 0.6211 and 0.5901 to 0.7033 and 0.7343, respectively. Combined with the signal removal, we can boot StarDist’s accuracy and F1-Score from 0.6211 and 0.5901 to 0.7717 and 0.8427. We hypothesize that this improvement is related to the training dataset used to train the model, which is relatively small compared to our dataset.

**Figure 4.**
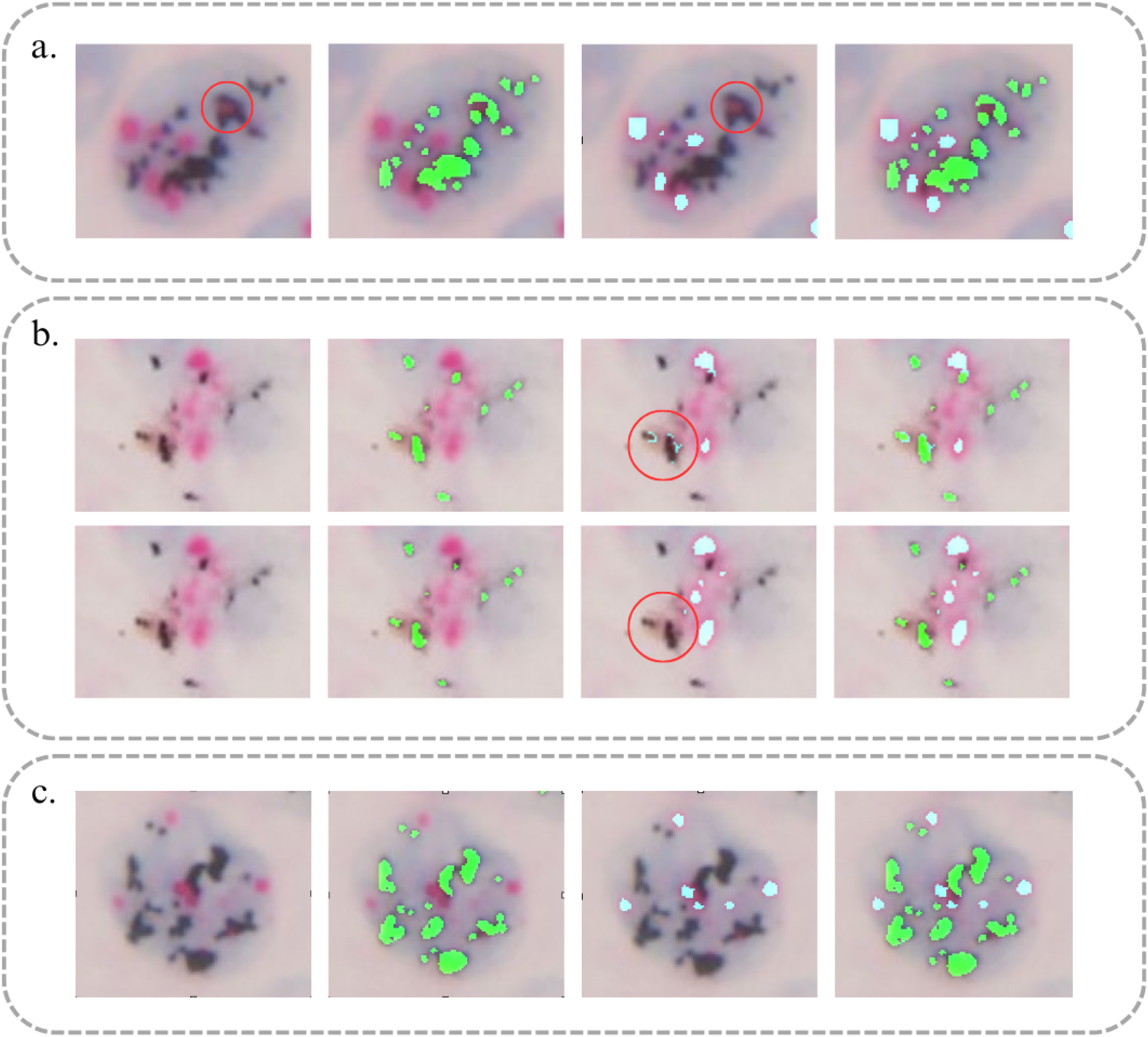
Cell boarder segmentation performance comparison. (a.) Overlapping of HER2 and Chr17 signals. (b.) One classifier for both signals versus Two classifier specialized for individual signals. (c.) Cell with HER2 overexpression.

### Same with the previous pre-processing method limited improvement was seen on Cellpose as parameters such as diameter can effectively allow Cellpose model to detect different sizes of cells

In general, the signal removal pre-processing method we proposed in this study can effectively improve the performance of StarDist and Cellpose model on DISH datasets, and resizing method have larger impact on StarDist model but have no benefit for the Cellpose model due to its ability to custom change the diameter of detecting object. The system is compatible with both models, allowing users to choose whether to apply this pre-processing treatment based on the specific model they intend to use for segmentation.

### Comparison of StarDist model with Cellpose model

We investigate the models that will be used to segment cell boarders for users to determine which will be more accurate, and more versatile. For this comparison, we selected the best-performing methods for each model: for StarDist-Both and Cellpose-Dug. Our findings indicate that Cellpose-Dug achieved slightly better results, with average accuracy and F1 scores of 0.8043 and 0.8608, respectively, compared to the StarDist-Both model’s 0.7717 and 0.8427. Cellpose also demonstrated lower standard deviations for both metrics, at 11.9 and 12.3, compared to StarDist’s 17.8 and 18.7 (4).

### Cell Feature Ranking Methods

The most straightforward way to calculate the DISH score is to determine the ratio of HER2 to Chr17 in each cell, then select the 20 highest-scoring cells and compute the average score. However, this score might be biased due to several reasons. First, the cells with the highest Her2/Chr17 ratio often have only a single HER2 signal, as evidenced by the average difference in Chr17 per cell, which is 1.236 higher in the manual readings compared to the auto-quantification system. Second is that cells with high HER2 to Chr17 don’t not necessarily exhibit morphology such as but not limited to abnormally large cell, elongated cell, and signals too close to the boarders, characteristics that pathologists deem as an appropriate candidate for the final analysis.

First, we select parameters such as area and sphericity of the cell body which is to mimic when two cells that have the same amount of HER2 and Chr17 signals, pathologist will prefer the one with more spherical and average in size (Eq. 1-2). Second, we also score the amount of Chr17 signals to prevent the system’s bias towards selecting cell with single Chr17 signals, to not leave out the cells with Chr17 signals but raise the priority of cells with two or more Chr17 signals (Eq. 3-5). Last, we normalize all the scores and add weightings for each score to control the importances of each characteristic in the final cell score.

We evaluated the best-performing weighting of the ranking equation (Eq. 6) against a simpler approach that ranks the ROI of the cells solely by their ratios in descending order. The performance of both models was compared with the pathologists’ manual readings. Without the ranking system, the overall percentage agreement (OPA) was 34.4%. In contrast, the best-performing ranking system achieved OPA values of 88.9%, respectively (1).

**Table 1.** Comparison of manual and automatic quantification results with and without ranking.

| Manual Result | Auto Quantification Results |  |  | Agreement |  |  |
| --- | --- | --- | --- | --- | --- | --- |
|  | Amplified | Non-amplified | Total | Measurement | Percentage | (n/N) |
| **without ranking** |  |  |  |  |  |  |
| Amplified | 22 | 0 | 22 | PPA | 100.0% | (22/22) |
| Non-amplified | 59 | 9 | 68 | NPA | 13.2% | (9/68) |
| Total | 18 | 72 | 90 | OPA | 34.4% | (31/90) |
| **with ranking** |  |  |  |  |  |  |
| Amplified | 17 | 5 | 22 | PPA | 77.3% | (17/22) |
| Non-amplified | 5 | 63 | 68 | NPA | 92.6% | (63/68) |
| Total | 22 | 68 | 90 | OPA | 88.9% | (80/90) |

### Comparison of Auto-quantification to manual pathologists’ results

We first compared our auto-quantification system with the ranking system the HER2 ratio, average HER2 signals, and average Chr17 signals across all 90 cases with the pathologists’ assessments. We compared agreement rates between the manual pathologist results and the auto-quantification system presented in this study. The resulting case-wise positive percentage agreement (PPA), negative percentage agreement (NPA), and overall percentage agreement (OPA) were 77.3%, 92.6%, and 88.9%, respectively (1).

**Figure 5.**
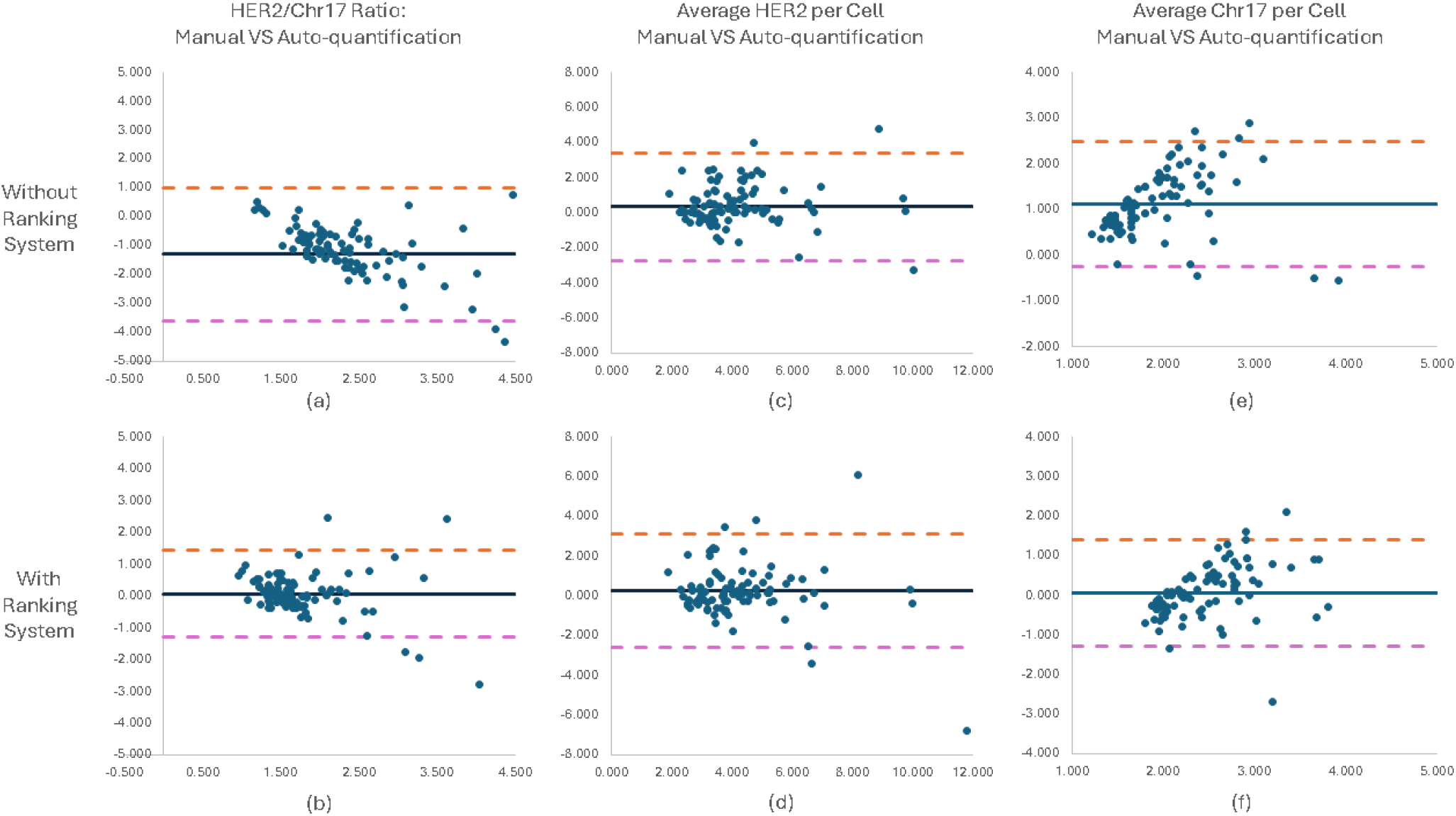
Evaluation of manual analysis VS Auto-quantification system using Bland-Altman plots. (a) (b) The comparison of manual analysis with auto-quantification system in terms of HER2/Chr17 ratio. (c) (d) Same comparison in terms of Average HER2 per cell. (e) (f) Same comparison in terms of Chr17 per cell.

Next, we confirmed the accuracy of the proposed system using Bland-Altman plots, the plot shows BIAS (blue), upper limit of agreement (orange), and lower limit of agreement (purple) (5). (5) shows the comparison of HER2/Chr17 ratio between manual DISH analysis and the auto-quantification system, the bias is 0.079 and the limit of agreement is −1.451 and 1.303. The comparison of average HER2, the bias is 0.246 and the limit of agreement is 3.109 and −2.617. Small bias and narrow limit of agreement of the HER2/Chr17 ratios shows that the system exhibits high accuracy in quantifying the HER2 amplification status.

The auto-quantification system demonstrated higher agreement in NPA, which is consistent with the finding of a lower average HER2 and a higher average Chr17 compared to the manual results. The higher disagreement rates observed in individual image cases were primarily due to instances where HER2 signals were elevated in one image but normal in another, leading to discrepancies depending on which image the pathologists selected for assessment. To address this, we offer a manual process allowing pathologists to discard undesirable cells, enabling more flexible adjustments and facilitating agreement between the pathologist and the auto-quantification system.

## Discussion

Based on our results, we have successfully introduced a system that is statistically proven to assist pathologists in assessing HER2 status in DISH images. The system will significantly reduce the labor and time intensive cost of HER2 analysis, and the lightweight design makes it assessable to low spec PC and open sources for all hospitals and pathology departments. Currently the system relies on HER2-DISH images that have an adequate contrast between the cell and the background to perform accurate boarder detection.

We acknowledge that variations in DISH images are likely to occur as new pathologists take charge of sample preparation. While we provide solutions for continued training of the classifier should pathologists find the existing classifier inadequate for analysis, the versatility of the Cellpose and StarDist models offers robust capabilities for capturing diverse cell border styles in future DISH images. We also observed that there are many cells with unwanted morphology such as but not limited to elongated cells, abnormally large and small cells, cells with too faint of both signals, and cells without HER2 and Chr17 cells. The ranking system alongside machine and deep learning model can easily rule out the unwanted cells, which can significantly speed up analysis time.

The limitation to this work is the cell ranking system as it still has room to tweak the weighting for each score. This work is like what a classifier can do; thus, we propose the inclusion of a classifier that takes in including the image of the cell, sphericity, etc. The classifier could ultimately mimic pathologist chose on which 20 cell to select narrowing differences of results between manual and auto-quantification.

Another limitation of this work includes being unable to perform boarder segmentation on HER2-DISH images while the cell border signals are inseparable to the background, as it is the curial step to perform the quantification task. We have observed about 5 in 120 cases that had this issue which was manually ruled out from our work. We propose the inclusion of an unsupervised learning model, such as CycleGAN, which can transform the style of an image to a predefined set style. The implementation of such a model could address the challenges we encountered with the classifier. By testing the performance of the best-performing model on the transformed style, we could potentially improve the accuracy of cell border segmentation and reduce the high variance observed in cell border segmentation.

Overall, the HER2 DISH auto-quantification system delivers a lightweight, open source, fast, and versatile tool for HER2 DISH analysis. The system fully automates the analysis process from HER2 and Chr17 segmentation by machine learning model, cell body segmentation by deep learning model, and a ranking system that helps pick out the most suitable cells for the 20 cells required in the HER2 DISH analysis. Pairing up with a user interface which allows pathologists to make manual corrections, which the whole analysis can be done within 1 to 2 minutes a comparable speed up to manual analysis.

## Methods and Materials

### 0.1 Sample Preparation

Current practice follows the procedure recommended by Roche, breast cancer tissue samples are fixed in 10% neutral buffered formalin are paraffin-embedded, sectioned to 4 micrometers, and mounted on Superfrost Plus slides. The HER2 and chromosome 17 probes, part of the VENTANA HER2 Dual ISH DNA Probe Cocktail, are hybridized to their respective target sequences during a dual in situ hybridization (ISH) process. Following hybridization, enzymatic reactions result in silver deposition for HER2 and red chromogen for Chr17. After counterstaining with hematoxylin, the slides are dehydrated, mounted, and scanned for digital imaging results. With different prob and slight changes in sample preparation such as but not limited to tissue obtain by core needle biopsy or by open biopsy, the size of the specimen, and the time treated by formalin might all lead to drastically different DISH image (1).

### 0.2 Pre-processing for Segmentation

Pre-processing plays a crucial role in the effectiveness of the segmentation processes in DISH images as there are HER2 and Chr17 signals that often effect the performance of the model, either mistaking the signal as a cell since both signals often are similar spherical shape with the cell or mistaking the signal for the cell boarder. For all methods of the cell boarder segmentation, we first convert the original DISH image into gray scale image, next remove both the HER2 and the Chr17 signals from the image with the mask we obtain from the previous step replacing the space with the average RGB value of the cell body (2). As for the StarDist model we observed that the re-scaling of the image drastically increases the model’s efficiency and accuracy.

### 0.3 Cell Boarder Segmentation

Upon segmenting the cell borders, the pre-processed image was passed through the selected model, which generated the detected borders. To evaluate segmentation performance, we manually created cell body ground truth for 20 DISH images sampled from a total of 316 DISH images. We then cropped the regions of interest (ROI) from 20 to 40 cells selected by our auto-quantification system, where HER2 status in agreement with the pathologists’ manual assessments.

We investigated three different segmentation approaches in combination with various pre-processing methods, resulting in a total of eight distinct methodologies. Initially, we compared the performance of classical algorithmic methods to the preferred deep learning methods. The classical algorithmic approach involved a combination of find edge filter, threshold, hole filling, and watershed algorithms, all implemented in ImageJ Fiji. Subsequently, we compared pre-processing techniques including a control (no signal removal), signal removal, and signal removal combined with resizing the image by a factor of 0.33 on both axes. This comparison revealed a significant performance enhancement with the application of our pre-processing method. Additionally, we assessed the performance of the StarDist model against the Cellpose model, enabling us to distinguish the differences between these two deep learning-based cell segmentation approaches. Finally, all comparisons of method performance on individual ROIs were conducted using the metrics of accuracy, precision, recall, and F1 score.

### 0.4 Ranking Cell ROI

For ranking cells for the final selection, we acknowledge that the guidelines suggest that the inspector chooses cell closer to the average size of the cells, while also cell must be representative cells of such cluster, such that we implemented and scoring system for individual cells based on parameter such as but not limited to area score, sphericity score, HER2 signal score, Chr17 signal score, signal ratio score (Eq. 1–6). All five scores of which will be given a weighting which all sums up to giving cells its rating. With this ranking system, we will test it against simply sorting the cells by individual cell’s ratios by their correlation and other metrics.

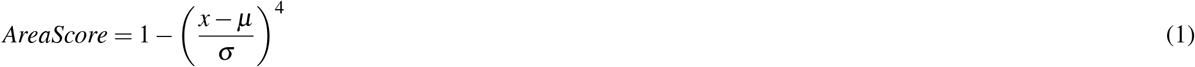

*x* = cell area, *µ* = average of cell area, *σ* = standard deviation of cell areas.

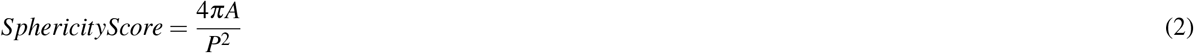

*A* = cell area, *P* = average of cell area.

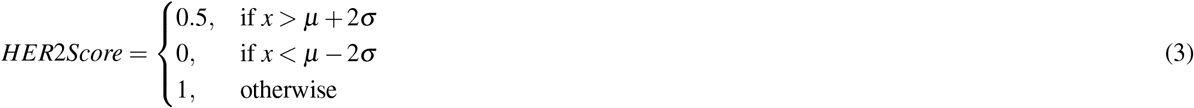

*x* = HER2 signal, *µ* = average HER2 signal, *σ* = standard deviation of HER2 signal.

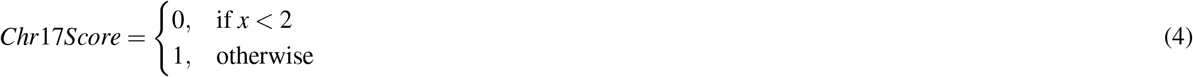

*x* = Chr17 signal.

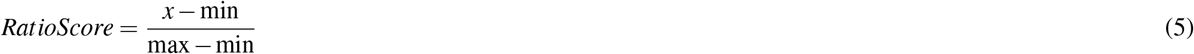

*min* = minimum value of 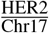ratio, *max* = maximum value of 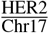ratio.

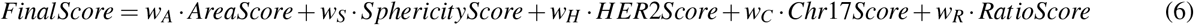

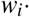 =weighting of each score.

### 0.5 Accuracy Evaluation

To validate the accuracy of our auto-quantification system, we compared its performance against the pathologists’ assessments across 90 different cases. We evaluated the system’s performance by analyzing true positive (TP), true negative (TN), false positive (FP), and false negative (FN) rates, along with positive percentage agreement (PPA), negative percentage agreement (NPA), and overall percentage agreement (OPA). These metrics demonstrate the system’s effectiveness in assisting pathologists with DISH image assessment.

## Data Availability

All data produced in the present study are available upon reasonable request to the authors

## Data availability

Due to privacy protections for patient histopathology image data and patient clinical data, we are unable to make all data public.

## Code availability

The data, source code, and standalone auto-quantification system for this study is available in HER2DISH and can be accessed via this link https://github.com/IanHuangOwO/HER2DISH.

## Acknowledgments

We gratefully acknowledge the support provided by Taoyuan General Hospital. Ministry of Health and Welfare and National Tsing Hua University for their invaluable assistance throughout this research.

## Author information

### Contribution

J.R.L. conceived the experiment(s) and setup prototype of the project. C.Y.H. conducted the experiment(s) finalized technical details, analyzed the results, and draft this manuscript. L.A.C. conceived the experiment(s), supervised the experiment(s) conducted by J.R.L. and C.Y.H. analyzed the results. P.C.H., C.H.L., and C.C.Y. provides professional opinion and assessment to the results. All authors reviewed the manuscript.

